# Prevalence of metabolic syndrome among patients with type 2 diabetes mellitus: A cross-sectional study in a tertiary care center of western Nepal

**DOI:** 10.1101/2024.06.03.24308403

**Authors:** Manoj Karki, Rejma Shrestha, Milan Dhungana, Bidhata Rayamajhi

## Abstract

**INTRODUCTION:** The coexistence of Metabolic Syndrome in Type 2 diabetic patients significantly increases the risk of stroke and cardiovascular disease. Due to its remarkably high prevalence, metabolic syndrome has gained significant interest over the last decade. Limited information exists regarding the occurrence of Metabolic Syndrome in Type 2 diabetic patients in developing nations like Nepal. Our study aims to determine the prevalence of Metabolic Syndrome among patients with Type 2 Diabetes Mellitus in Western Nepal.

**METHODS:** This prospective cross-sectional study was conducted at the Universal College of Medical Sciences (UCMS) among patients with Type 2 Diabetes Mellitus. Patients with gestational diabetes mellitus, Type I Diabetes Mellitus, and those aged less than 20 were excluded. Metabolic syndrome has been diagnosed based on the definition given by the International Diabetes Federation.

**RESULTS:** The study involved 123 patients with a mean age of 61.44 ± 12.88 years, predominantly female (55.3%). Of these patients, 42.3% were classified as Obese. The overall prevalence of Metabolic Syndrome in Type 2 Diabetes Mellitus patients was 61%, comprising 75 cases. The most common component in our study was hypertension, with 82 cases (66.7%), followed by central obesity with 81 cases (65.9%). Among patients with Metabolic Syndrome, 66.7% had an estimated glomerular filtration rate (eGFR) in the 60-89 ml/min range. In females, central obesity was the most common component, while in males, hypertension was the most common component of Metabolic Syndrome.

**CONCLUSION:** The prevalence of Metabolic Syndrome in patients with Type 2 Diabetes Mellitus was high. Therefore, timely detection and management of Metabolic Syndrome is crucial in preventing consequences and premature mortality in individuals with Type 2 Diabetes Mellitus.

## Introduction

Metabolic Syndrome (MetS) is a complex disorder characterized by central obesity, hypertension, hyperglycemia, decreased HDL cholesterol, and elevated triglycerides. These conditions increase the risk of atherosclerotic cardiovascular diseases and Type 2 Diabetes Mellitus (T2DM). The global prevalence of MetS has risen from 37.6% in 2011-12 to 41.8% in 2017-18, affecting 20-30% of adults [1,2].

In Pakistan, MetS prevalence is 34.8% (IDF criteria) and 49% (modified NCEP ATP III criteria). India shows regional disparities: 31.4% in eastern, 29.7% in southern, and 5.0% in central regions, with higher rates among females (48.2%) than males (16.3%) [3]. In Nepal, a survey indicated a prevalence of 15-16% (NCEP ATP III and IDF criteria). Eastern Nepal reported a 22.5% prevalence (IDF criteria) and 20.7% (NCEP definition), suggesting a lower burden than other South Asian nations [4].

MetS significantly increases the risk of T2DM and cardiovascular diseases, with a three to five-fold higher risk of T2DM. Among individuals with T2DM, MetS prevalence ranges from 45.8% to 96.3% [5]. Future projections indicate a substantial rise in diabetes cases, particularly in developing nations, with over 70% of new cases expected [6]. The IDF Diabetes Atlas predicts the number of adults with diabetes in Southeast Asia will reach 113 million by 2030 and 151 million by 2045 [7], with 541 million adults currently having impaired glucose tolerance [8].

T2DM is a significant health concern in South Asia, with an expected 151% increase in prevalence from 2000 to 2030 [9]. Countries like India and China are among the top ten with the highest number of diabetes patients. A study in Nepal reported an 8.5% prevalence among adults with T2DM [10]. The severe impact of MetS in T2DM patients reduces survival rates by at least ten years, highlighting the need for aggressive risk factor management by primary care physicians. Understanding MetS components in western Nepal can inform effective intervention and prevention strategies.

## Methods

### Study Design

This hospital-based descriptive, cross-sectional study was conducted in the Department of Internal Medicine at Universal College of Medical Sciences Teaching Hospital (UCMSTH) from September 01, 2021, to February 28, 2023, using a purposive non-probability sampling technique. Institutional approval was obtained from the Institutional Review Committee (IRC) of UCMS (UCMS/IRC/134/21). The study included patients with Type 2 Diabetes Mellitus attending the inpatient and outpatient departments of Internal Medicine. Exclusion criteria were patients with Type 2 Diabetes Mellitus under 20 years of age, those with gestational diabetes, and those with Type 1 Diabetes Mellitus.

### Sampling and Statistical Analysis

Convenience sampling was done, and the sample size was calculated using the formula.

***n = z2 pq/d2***

where,

z = 1.96 taken at a 95 % of confidence interval

n = Required sample size

p = Prevalence of Metabolic Syndrome in patients with Type 2 Diabetes Mellitus (66.2%)

q = 100 – p

d = 8.5 % (Maximum tolerable error)

Based on the above formula, the estimated sample size at a 95% confidence interval and 8.5% error was 118; hence, the total sample collected was 123.

The study employed a comprehensive approach to gather patient data, encompassing clinical history, socioeconomic demographics, and diagnostic criteria outlined by the internationally recognized International Diabetes Federation for Metabolic Syndrome [11]. Informed written consent was obtained from each participant. Each participant voluntarily consented to participate in this research and understands the details, purposes, and possible future implications of the study and their right to withdraw at any time.

Tools included a structured questionnaire (Performa) for patient information like age, sex, history, physical examination, presence of comorbidities, socioeconomic status, clinical findings like blood pressure and anthropometric measurements, and baseline investigations such as complete blood count, liver and thyroid function tests, lipid profile, fasting and postprandial blood glucose, ECG, urine analysis, renal function tests, and eGFR calculation.

Data analysis utilized SPSS version 17, where frequencies, percentages, means, standard deviations, and p-values were computed to analyze the collected data comprehensively.

## Results

In this study involving 123 patients, the mean age was 61.44 ± 12.88 years. The largest age group was 60-69, comprising 28.5% (35 patients). The next largest group was 50-59 years old, accounting for 25.2% (31 patients). There was a decline in the number of younger patients: 13% (16 patients) were 40-49 years old, 10.6% (3 patients) were 30-39 years old, and only 2.4% (2 patients) were 20-29 years old. Among older patients, 18.7% (23 patients) were 70-79 years old, and 10.6% (13 patients) were 80-89 [Table 1].

**Table 1:** Baseline characteristics.

| **Gender** | **Number** | **Age in Years (Mean ± sd)** |
| --- | --- | --- |
| Male | 55 (44.7%) | 60.93± 12.22 |
| Female | 68 (55.3%) | 63.79± 10.66 |
| **Age groups (Years)** | **Number according to age group** |  |
| 20-29 | 2 |  |
| 30-39 | 3 |  |
| 40-49 | 16 |  |
| 50-59 | 31 |  |
| 60-69 | 35 |  |
| 70-79 | 23 |  |
| 80-89 | 13 |  |
| **BMI Category** | **Percentage (%)** |  |
| Underweight | 2.4 |  |
| Normal | 18.7 |  |
| Overweight | 23.6 |  |
| Obesity I | 42.3 |  |
| Obesity II | 13 |  |
| **Duration of onset of diabetes** | **Number (%)** |  |
| <10 years | 101 (82.1%) |  |
| 11-20 Years | 16 (13%) |  |
| >20 years | 6 (4.9%) |  |

The mean age for male patients was 60.93 years with a standard deviation of 12.22 years, while for female patients, the mean age was 63.79 years with a standard deviation of 10.66 years. Of the total, 55 were male (44.7%) and 68 were female (55.3%) [Table 1].

Regarding BMI distribution, the overall mean BMI was 25.85±4.54. Regarding BMI categories, 42.3% of patients fell into the Obese I category, 23.6% were Overweight, 18.7% had a Normal BMI, 13.0% had Grade II obesity, and 2.4% were Underweight [Table 1].

When examining the duration of type 2 diabetes mellitus among the patients, 82.1% (101 cases) had been living with the condition for less than ten years. Additionally, 13% (16 cases) had diabetes for 11-20 years, and 4.9% (6 cases) had been managing diabetes for more than 20 years [Table 1].

Out of the total 123 cases, 75 individuals (61%) were diagnosed with Metabolic Syndrome. The overall percentage prevalence of Metabolic Syndrome in Type 2 Diabetes Mellitus patients was 61%, comprising 75 cases.

### Prevalence of Metabolic Syndrome according to sex

Among the 55 male cases, 29 individuals (52.7%) and among the 68 female cases, 46 individuals (67.6%) were diagnosed with Metabolic Syndrome. The prevalence of Metabolic Syndrome in Type 2 Diabetes Mellitus is 52.7% and 67.6% in males and females, respectively [Table 2].

**Table 2:** Table showing the Prevalence of Metabolic Syndrome according to sex.

| Sex of the patient | Metabolic Syndrome |  | p-value |
| --- | --- | --- | --- |
|  | No | Yes |  |
| Male | 26(47.3) | 29(52.7) | 0.092 |
| Female | 22(32.4) | 46(67.6) |  |

### Association between gender and components of metabolic syndrome

In females, central obesity was the most common component, consisting of 50 cases, followed by hypertension (41 cases), then raised triglyceride (32 cases), and the least common component was reduced HDL-Cholesterol comprising of 27 cases. In males, hypertension was the most common component, comprising 41 cases, followed by central obesity, comprising 31 cases; then reduced HDL-Cholesterol, comprising 29 cases, and the least common component was raised triglyceride, comprising 18 cases. There was a significant association between central obesity and the gender of the patient with Metabolic Syndrome. Central obesity was higher in females than males (p-value = 0.046). However, other components of metabolic syndrome were not significant, based on the gender of the patients [Table 3].

**Table 3:** Cross tabulation shows an association between gender and components of metabolic syndrome in study patients.

| Components of Metabolic Syndrome |  | Sex of the patient |  | P value |
| --- | --- | --- | --- | --- |
|  |  | Male | Female |  |
| Central Obesity | No | 24 (57.1%) | 18 (42.9%) | 0.046 |
|  | Yes | 31 (38.3%) | 50 (61.7%) |  |
| Raised Triglycerides | No | 37 (50.7%) | 36 (49.3%) | 0.108 |
|  | Yes | 18 (36%) | 32 (64%) |  |
| Reduced HDL-cholesterol | No | 26 (38.8%) | 41 (61.2%) | 0.149 |
|  | Yes | 29 (51.8%) | 27 (48.2%) |  |
| Hypertension | No | 14 (34.1%) | 27 (65.9%) | 0.096 |
|  | Yes | 41 (50%) | 41 (50%) |  |

### History of alcohol consumption

93 cases (75.6 %) had a history of alcohol consumption while the remaining 30 cases (24.4%) did not have a history of alcohol consumption.

### History of smoking habit

36 cases (29.3%) were smokers, and the remaining 87 cases (70.7%) were non-smokers.

### eGFR of the patient with metabolic syndrome

Among 75 patients with Metabolic Syndrome, 22 males had an eGFR of 60-89 ml/min, 3 had 45-59 ml/min, and 2 each had 30-44 ml/min and >90 ml/min. For females, 28 had an eGFR of 60-89 ml/min, 6 had 45-59 ml/min, 4 had 30-44 ml/min, 3 had 15-29 ml/min, and 3 had >90 ml/min, with 2 below 15 ml/min. Kidney function impairment was observed in 27 males (39%) and 43 females (61%) [Fig 1].

**Figure 1:**
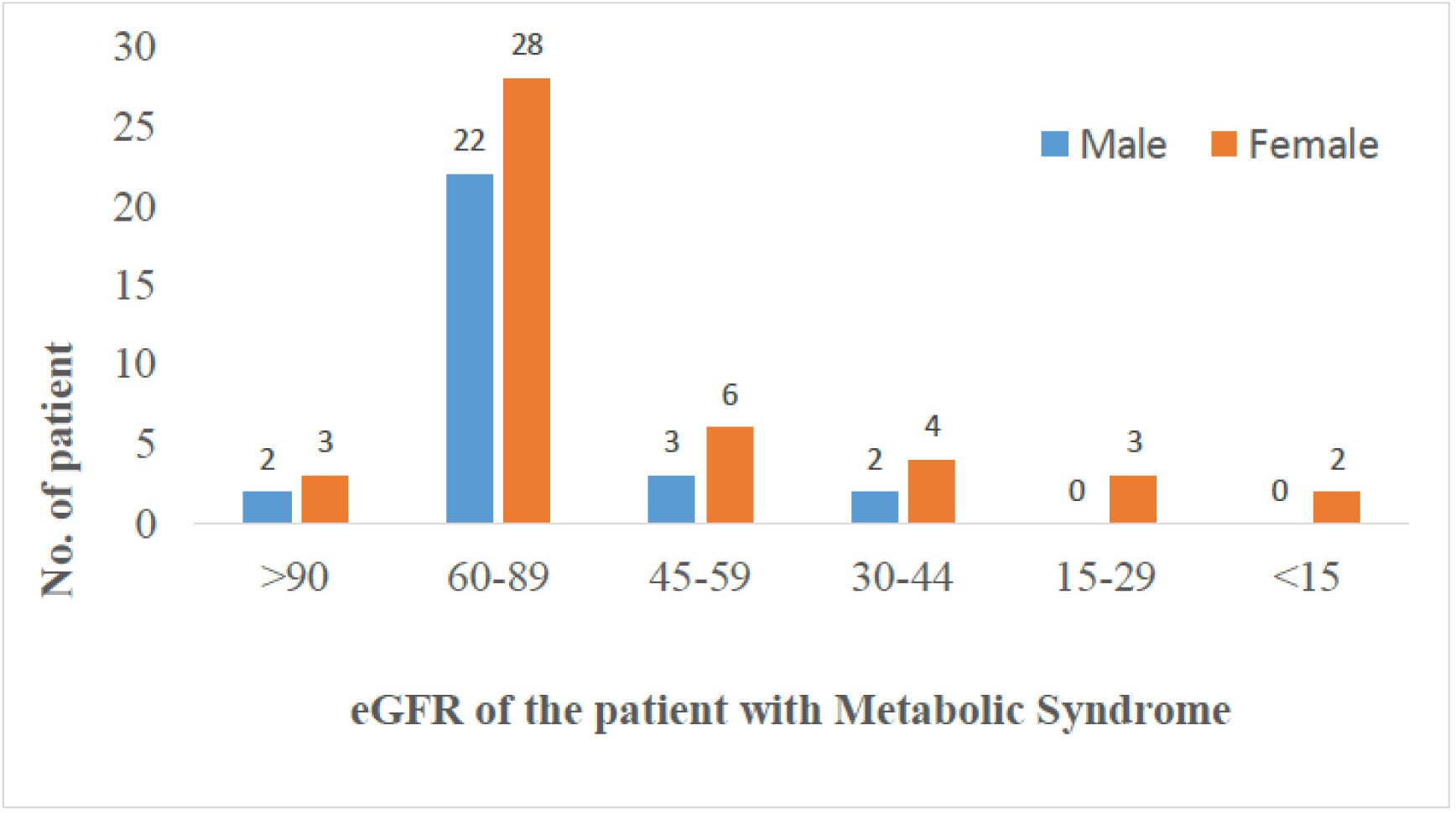
Distribution of eGFR of the patient with metabolic syndrome according to gender (N= 75).

## Discussion

In our study, the mean age for Males was 60.93 years, with a standard deviation (SD) of 12.22 years, and females was 63.79 years, with an SD of 10.66 years, which is like a study conducted by Yeboah O et al. [12] in Ghana, where the mean age was 56.42 ± 10.64. In our study, the prevalence gradually increased with the increase in age and remained highest in the age range of 60–69 years, which aligns with the study conducted by Pokhrel R et al. [13] in Pokhara, Nepal, in the age range of 50– 69 years.

In our study, females had a higher prevalence of metabolic syndrome at 67.6% compared to males with a prevalence of 52.7%, like studies conducted by Yeboah O et al. [12], where the prevalence in females was 62.35%. A similar conclusive finding was obtained from the study conducted by Pokhrel R et al. [13] in Pokhara, Nepal, where the prevalence among female patients was much higher. A study by Efaishat R et al. [14] stated that there was no significant difference in the prevalence of metabolic syndrome among the sexes. In sharp contrast to the current study’s findings, metabolic syndrome was higher among men than women in a study conducted by Puppet F et al. [15] in Urban North-Central Nigeria, where the prevalence of males was 74.5%. The authors justified that the high activity profile of women might contribute to the exceptionally higher prevalence of metabolic syndrome among men in Urban North-Central Nigeria.

In our study, the mean BMI of patients with Type 2 Diabetes Mellitus was 25.85 kg/m2, with a standard deviation of 4.54, which is consistent with a study conducted by Nsiah K et al. [16] with the average mean BMI value was 26.43 kg/m2. The highest duration of Type 2 Diabetes Mellitus since its onset was 1-10 years, accounting for 101 cases (82.1%). In contrast, in a study conducted by Wube B et al. [17], 55.7% of participants had Type 2 Diabetes Mellitus for less than five years. This finding aligns with the study conducted by Pokhrel R et al. [13] in Pokhara, Nepal, where the mean duration of type 2 diabetes mellitus was six years.

In our study, the prevalence of Metabolic Syndrome in patients with Type 2 Diabetes Mellitus was found to be 61%, comprising 75 cases. Prevalence in males and females was 52.7% and 67.6% respectively. This figure aligns with other studies conducted in Nepal using the IDF criteria: In Chitwan, Sharma K et al. [18] found a prevalence of 58.4%, while in Pokhara, Pokhrel R et al. [13] reported a prevalence of 66.8%. Additionally, in Ghana, Yeboah O et [12] observed a prevalence of 69.14%, and Wube B et al. [17] reported a prevalence of 59.9%. Our study findings were consistent with other research that assessed the prevalence of Metabolic Syndrome in Type 2 Diabetes Mellitus using the NCEP/ATP III criteria: Gemeda D et al. [19] reported a prevalence of 68.3%, and Nsiah K et al. [16] found a prevalence of 58%, Uprety T et al. [20] Sanepa, Nepal, reported a prevalence of 68.5%. A study conducted by Yeboah O. et al. concluded that the prevalence and principal components of the syndrome vary among populations due to the influence of genetic and lifestyle factors [12]. It indicates that the prevalence of Metabolic Syndrome is high among diabetes patients, which is expected because they were already suffering from Type 2 Diabetes Mellitus, which itself is an entity of the Metabolic Syndrome. Whereas the prevalence was even higher in the studies conducted by Bhatti G. et al. [21] in Urban India (71.9%), Yeboah F et al. [22] in Kumasi, Ghana (90.6%), and Kengne A et al. [11] in Cameroon (71.7%). This discrepancy could be due to the differences in the study period and sample size, as well as the level of hospital and population characteristics.

In our study, the most common component was hypertension and central obesity, which comprised 82 cases (66.7%) and 81 cases (65.9%), respectively. It was then followed by reduced HDL-cholesterol comprising 56 cases (45.5%). The least common component in our study was raised triglyceride, found in 50 cases (40.7%). These findings align with the findings conducted by Nsiah K et al. [16], where hypertension was the commonest component of Metabolic Syndrome, comprising 60%, followed by central obesity. However, our finding does not correlate with the study conducted by Pokhrel R et al. [13] in Pokhara, Nepal, where hypertension is the least common component. The discrepancy in the prevalence of metabolic syndrome between these studies could be due to differences in ethnic or geographical areas. Research in the past shows central obesity is the most prevalent component of Metabolic Syndrome in Nepal [23]

In our study, central obesity was found more in females than in males (p—value=0.046), which is consistent with the findings conducted by Herath M et al. [24] Srilanka. Pandit K et al. [25] in Kolkata, India, concluded that abdominal obesity was quite prevalent, with females outnumbering males in South Asian countries.

In our study, out of the total 75 cases who had Metabolic Syndrome, 50 cases comprising 66.7% had an eGFR in the range of 60-89 ml/min, falling into the category of mildly reduced eGFR, defined as a value between 60 and 90 mL/min/1.73 m2. This finding is consistent with the study conducted by Hu W et al. [26] China concluded that Metabolic Syndrome is associated with an increased risk of a mildly reduced eGFR in the Chinese population, and several individual components of Metabolic Syndrome have different impacts on eGFR levels: elevated TG, reduced HDL, and obesity increased risk for a mildly reduced eGFR. Hence, Metabolic Syndrome has dual roles in renal damage.

Kidney function impairment in patients with Metabolic Syndrome was observed in 27 male individuals (39%), while the remaining 43 were females, accounting for 61%. This finding is inconsistent with the study conducted by Song H et al. [27], where Men had a higher prevalence of reduced eGFR than women. The discrepancy could be attributed to the fact that our study group had a higher proportion of females (55.3%) than Song H et al. (43.7%).

In our study, out of the total 75 cases of metabolic syndrome, 20 cases, comprising 26.7%, had an eGFR of less than 60 ml/min, falling into the category of chronic kidney disease. This finding is inconsistent with the study conducted by Maleki A. et al. [28] in Iran and Zhang L. et al. [29] in China, where the prevalence of CKD in patients with metabolic syndrome is 14.8% and 15.4%, respectively. This difference can be attributed to the distinct study populations; our study focused on patients with Type 2 Diabetes Mellitus, a significant cause of CKD, whereas the other studies examined the general population.

This study has certain limitations. This study, conducted within a single center and a limited timeframe, focused on patients with type 2 diabetes mellitus, significantly influencing glycemic control, weight, and waist-hip ratio. The small sample size, reflective of a hospital-based setting, limits the representativeness and generalizability of the findings, particularly given the urban-centric utilization of health services in western Nepal. Additionally, the cross-sectional design precluded follow-up on patient outcomes and the establishment of cause-effect relationships, and the absence of a control group further complicates the generalization of the results.

## Conclusion

The study found that Type 2 Diabetes Mellitus patients had Metabolic Syndrome, with a higher prevalence in females compared to males. Hypertension was the most common component, followed by central obesity, low HDL-cholesterol, and high triglycerides. Central obesity was more common in females, while hypertension was predominant in males. Kidney function impairment was prevalent among these patients, with most experiencing mild impairment, and it was more common in females. Given the significant burden of Type 2 Diabetes Mellitus on individuals and healthcare systems, early detection and management of Metabolic Syndrome and its risk factors are crucial to prevent complications.

## Data Availability

Data analysis was done with the help of computer using Epidemiological Information. Using this software, frequencies, percentage, mean, standard deviation and'p' values were calculated. Data was collected, entered and analyzed in SPSS software version 17.

## Acknowledgment

The authors would like to thank the Department of Internal Medicine, Universal College of Medical Sciences, and all the participants involved in this study for their assistance in the research project.

